# Analytical assessment of Beckman Coulter Access anti-SARS-CoV-2 IgG immunoassay

**DOI:** 10.1101/2020.11.05.20226555

**Authors:** Maurizio Ruscio, Elisa D’Agnolo, Anna Belgrano, Mario Plebani, Giuseppe Lippi

## Abstract

**Background:** The approach to diagnosing, treating and monitoring severe acute respiratory syndrome coronavirus 2 (SARS-CoV-2) infection relies strongly on laboratory resources, with serological testing representing the mainstay for studying the onset, nature and persistence of humoral immune response. This study was aimed at evaluating the analytical performance of the novel Beckman Coulter anti-SARS-CoV-2 IgG chemiluminescent immunoassay.

**Methods:** This analytical assessment encompassed the calculation of intra-assay, inter-assay and total imprecision, linearity, limit of blank (LOB), limit of detection (LOD), functional sensitivity, and comparison of anti-SARS-CoV-2 antibodies values obtained on paired serum samples using DiaSorin Liaison SARS-CoV-2 S1/S2 IgG and Roche Elecsys Anti-SARS-CoV-2 total antibodies. Diagnostic performance was also tested against results of molecular testing on nasopharyngeal swabs, collected over the previous 4 months.

**Results:** Intra-assay, inter-assay and total imprecision of Beckman Coulter anti-SARS-CoV-2 IgG were between 4.3-4.8%, 2.3-3.9% and 4.9-6.2%, respectively. The linearity of the assay was excellent between 0.11-18.8 antibody titers. The LOB, LOD and functional sensitivity were 0.02, 0.02 and 0.05, respectively. The diagnostic accuracy (area under the curve; AUC) of Beckman Coulter anti-SARS-CoV-2 IgG compared to molecular testing was 0.87 (95% CI, 0.84-0.91; p<0.001) using manufacturer’s cut-off, and increased to 0.90 (95% CI, 0.86-0.94; p<0.001) with antibody titers. The AUC was non-significantly different from that of Roche Elecsys Anti-SARS-CoV-2, but was always higher than that of DiaSorin Liaison SARS-CoV-2 S1/S2 IgG. The correlation of Beckman Coulter Access SARS-CoV-2 IgG was 0.80 (95% CI, 0.75-0.84; p<0.001) with Roche Elecsys Anti-SARS-CoV-2 and 0.72 (95% CI, 0.66-0.77; p<0.001) with DiaSorin Liaison SARS-CoV-2 S1/S2 IgG, respectively.

**Conclusions:** The results of this analytical evaluation of Beckman Coulter Access anti-SARS-CoV-2 IgG suggests that this fully-automated chemiluminescent immunoassay represents a valuable resource for large and accurate seroprevalence surveys.

## Introduction

Coronavirus Disease 2019 (COVID-19) can now be considered the biggest tragedy that has affected humanity since the end of the Second World War, in 1945 (1). With over 1.3 million casualties so far and following an epidemic trajectory that is very unlikely to reverse soon, the ongoing severe acute respiratory distress syndrome coronavirus 2 (SARS-CoV-2) pandemic outbreak is deeply disrupting health care, economy and even social relationships in all worldwide countries (2).

It is now virtually unquestionable that the approach for diagnosing, treating and monitoring COVID-19 shall be strongly based on laboratory investigations, with three levels of in vitro diagnostic tests (3). Molecular or antigenic detection of SARS-CoV-2 represents the mainstay for diagnosing an acute infection, and thereby for isolating or treating infected and potentially infective patients (4). Routine hematological and biochemical testing is essential for defining disease severity and for eventually predicting illness progression (5), whilst serological testing, which can be defined as a diagnostic investigation used for revealing and monitoring the development of an immune response against a given pathogen (6), is essentially aimed at unraveling as to whether some subjects have been infected by SARS-CoV-2 and have then developed a humoral immune response, as reflected by production of different classes of antiviral immunoglobulins (IgG) (7). Serological testing can hence find its most rationale foundation within the context of seroprevalence studies, for assessing nature and extent of humoral immunity against SARS-CoV-2, for screening convalescent plasma, for monitoring herd immunity, either natural or consequent to widespread vaccination, as well as for supporting molecular testing in some well-defined circumstances (7).

The serological assessment of patients who currently have, or have developed, a certain or presumptive SARS-CoV-2 infection is extremely variegated, especially in terms of antibodies tested, analytical methods and turnaround time (8). Briefly, the immunoassays for detecting anti-SARS-CoV-2 antibodies have been developed against single immunoglobulin classes (i.e., IgA, IgM or IgG) or against total immunoglobulins, can be either based on laboratory-based or point of care (POC) techniques, and encompasses a vast array of analytical principles, spanning from rapid lateral flow immunoassays (LFIAs), manual enzymatic linked immunosorbent assay (ELISAs), to fully automated techniques such as chemiluminescence (CLIAs) or fluorescent (FIA) immunoassays (9,10).

Irrespective of the assay and its purpose, the Task Force on COVID-19 of the International Federation of Clinical Chemistry and Laboratory Medicine (IFCC) strongly advises that each method must be accurately evaluated and validated before being introduced into routine SARS-CoV-2 diagnostics (11). According to this clear-cut preamble, the main aim of this study was to evaluate the analytical performance of a novel anti-SARS-CoV-2 IgG immunoassay, recently developed and commercialized by Beckman Coulter.

## Materials and Methods

### Immunoassay description

The novel Beckman Coulter Access SARS-CoV-2 IgG (Beckman Coulter Inc., Brea, CA, USA) is a two-step CLIA. Briefly, 20 μL of patient sample are mixed with buffer within a reaction cuvette along with paramagnetic particles coated with recombinant SARS-CoV-2 S1 protein containing the amino acid sequence of the receptor binding domain (RBD). After incubation, the antibodies bound to the solid phase are sequestrated thorough generation of a magnetic field, whilst unbound material is eliminated by washing. An anti-human IgG alkaline phosphatase conjugate monoclonal antibody is then added, followed by second mixture separation by washing, for removing unbound conjugate material. A chemiluminescent substrate is finally pipetted into the cuvette, and the amount of light generated is measured using a luminometer. The amount of light produced is plotted versus the cut-off value calculated during assay calibration. Final results ≥1 are reported as being reactive for SARS-CoV-2 IgG antibodies, values between 0.8-1.0 as equivocal and those ≤0.80 as non-reactive for SARS-CoV-2 IgG antibodies. The calibration curve is stable for up to 28 days and the entire assay can be completed within 32 min.

### Method evaluation

The assessment of this novel Beckman Coulter Access SARS-CoV-2 IgG immunoassay on UniCel DxI800 has originally encompassed the calculation of intra-assay, inter-assay and total imprecision, linearity, limit of blank (LOB), limit of detection (LOD), functional sensitivity, as well as comparison of anti-SARS-CoV-2 antibodies values obtained on paired serum samples using two other commercial anti-SARS-CoV-2 immunoassays, as comprehensively described below. Serological testing was carried out using routine serum samples collected in 5 mL Vacutainer tubes (Greiner, with separation gel), from the healthcare personal working in the hospital of Trieste, Italy. All samples were centrifuged at 3500 rpm for 15 min at room temperature. Serum was then separated from the underneath cell layer, and 1 mL aliquot stored in 3 mL Criovial and frozen at – 20°C. At time of testing, the aliquot was allowed to thaw at room temperature, was then accurately mixed and used for serological assessment. An identical lot of reagents and the same calibration curves were used for all assays, throughout the study period.

### Imprecision

The intra-assay, inter-assay and total imprecision of Beckman Coulter Access SARS-CoV-2 IgG has been calculated using two serum samples displaying low (i.e., below the cut-off: ∼0.11) and high (i.e., above the cut-off: ∼8.33) high anti-SARS-CoV-2 IgG titers. Specifically, intra- and inter-assay imprecision were assayed in 10 consecutive replicate runs and 10 consecutive working days (duplicate measure every day), respectively. The imprecision of the assay has been finally calculated as coefficient of variation (CV%). Total imprecision has been was estimated according to the equation suggested by Krouwer and Rablnowitz (12).

### Linearity

The linearity of Beckman Coulter Access SARS-CoV-2 IgG has been assessed by measuring in duplicate serial dilutions (e.g., from 1:9 to 9:1) of a serum sample with high anti-SARS-CoV-2 IgG titer (i.e.,18.8) and a second serum sample with low anti-SARS-CoV-2 IgG titer (i.e., 0.11). The estimated and measured anti-SARS-CoV-2 IgG values were then correlated with linear fit, and with calculation of Spearman’s correlation.

### Limit of blank, limit of detection and functional sensitivity

The LOB and LOD were calculated using the formula [LOB] = [mean value] + [1.645]×[standard deviation, SD)] of 20 replicates of sample buffer, and [LOD] = [LOB] + [1.645]×[Standard Deviation] of 20 replicates of a serum sample with the lowest measurable anti-SARS-CoV-2 IgG titer, as suggested by Armbruster and Pry (13). The functional sensitivity of the method was calculated as the lowest anti-SARS-CoV-2 IgG measurable value with arbitrary imprecision set at ≤10%. Specifically, it was estimated with measurement of 8 different 1:2 scalar dilutions with sample buffer (from 1:1 to 1:128) of a routine serum sample with anti-SARS-CoV-2 IgG titer of 8.82. The scalar dilutions were then tested with 10 consecutive replicates, with calculation of imprecision obtained for each dilution. A model fit was developed for extrapolating the anti-SARS-CoV-2 IgG value which could be assayed with ≤10% imprecision.

### Diagnostic performance and immunoassays comparison

As previously mentioned, comparison studies were based on a seroprevalence survey carried out using on serum samples of healthcare professionals working at the hospital of Trieste, who also had a nasopharyngeal swab collected over the preceding 4 months. The results of Access SARS-CoV-2 IgG were then compared with those obtained on paired serum samples using DiaSorin Liaison SARS-CoV-2 S1/S2 IgG and Roche Elecsys Anti-SARS-CoV-2 total antibodies. The characteristics of the three immunoassays are summarized in table 1. The concordance of SARS-CoV-2 antibody titers obtained with the three methods was evaluated using Spearman’s correlation, whilst the agreement with molecular testing, and more precisely the area under the curve (AUC), diagnostic agreement, diagnostic sensitivity and diagnostic specificity, were calculated using Receiver Operating Characteristics (ROC) curve analyses. The diagnostic agreement with results of molecular testing was estimated with either continuous (i.e., antibody titer) or dichotomous (positive or negative results compared to manufacturers’ cut-offs) data. Molecular testing for detecting SARS-CoV-2 RNA on nasopharyngeal samples was carried out with Allplex 2019-nCoV Assay (BioRad, Basel, Switzerland), whose technical and analytical characteristic have been previously described elsewhere (14).

**Table 1.** Principal characteristics of the anti-SARS-CoV-2 chemiluminescent immunoassays used in this study.

| Manufacturer | Assay | Platform | Antibody class | Target antigen | Cut-off for positivity |
| --- | --- | --- | --- | --- | --- |
| Beckman Coulter, Brea, CA, USA | Access SARS-CoV-2 IgG | UniCel DxI800 | IgG | Spike protein (S1 – receptor binding domain) | $\geq 1$ |
| DiaSorin, Saluggia, Italy | Liaison SARS-CoV-2 S1/S2 IgG | Liaison XL | IgG | Spike protein (S1/S2) | $\geq 15$ |
| Roche Diagnostics, Indianapolis, IN, USA | Elecsys Anti-SARS-CoV-2 | Cobas 8000 | Total Ig | Nucleocapsid protein | $\geq 1$ |

### Statistical analysis

The statistical analysis was performed using Analyse-it (Analyse-it Software Ltd, Leeds, UK). The entire investigation was based on pre-existing serum samples, collected for routine SARS-CoV-2 testing during a hospital seroprevalence study, and thereby no patient’s informed consent or Ethical Committee approval were necessary.

## Results

### Imprecision

The results of imprecision study using two serum samples with low and high anti-SARS-CoV-2 IgG titers are shown in table 2. Briefly, the intra-assay, inter-assay and total imprecision were comprised between 4.3-4.8%, 2.3-3.9% and 4.9-6.2%, respectively.

**Table 2.** Intra-assay, inter-assay and total imprecision of Beckman Coulter Access SARS-CoV-2 IgG.

| Pools | Within-run (n=10) |  | Between-run (n=10) |  | Total |
| --- | --- | --- | --- | --- | --- |
|  | Mean±SD<br>(ng/L) | Imprecision<br>(CV %) | Mean±SD<br>(ng/L) | Imprecision<br>(CV %) | Imprecision<br>(CV %) |
| Low | 0.114±0.005 | 4.3% | 0.113±0.003 | 2.3% | 4.9% |
| High | 8.327±0.397 | 4.8% | 8.004±0.312 | 3.9% | 6.2% |

*CV, coefficient of variation; SD, standard deviation.*

### Linearity

The linearity of the novel Beckman Coulter Access SARS-CoV-2 IgG was found to be excellent over the range of anti-SARS-CoV-2 IgG titers tested. More specifically, the linearity (Spearman’s correlation) was found to be r=0.997 (p<0.001) between anti-SARS-CoV-2 IgG titers of 0.11 and 18.8.

### Limit of blank, limit of detection and functional sensitivity

The LOB, LOD and functional sensitivity (antibody titer), calculated according to the criteria previously mentioned, were found to be 0.02, 0.02 and 0.05, respectively. Notably, the latest sample dilution (i.e., 1:128), corresponding to anti-SARS-CoV-2 IgG antibody titer of 0.05, was associated with ∼6% imprecision. It is hence predictable that the functional sensitivity of this immunoassay could have been even lower than this value.

### Diagnostic performance and immunoassays comparison

The final sample size for diagnostic performance and immunoassays comparison studies consisted of 305 serum samples collected form hospital workers undergoing routine SARS-CoV-2 testing, for whom a definitive result (negative or positive) of molecular test on nasopharyngeal swab was available within the previous 4 months (range 0.5-4.0 months), which is the most suitable diagnostic window for detecting IgG humoral immunity against SARS-CoV-2 (15). The diagnostic performance of the three different immunoassays (i.e., Beckman Coulter Access SARS-CoV-2 IgG, DiaSorin Liaison SARS-CoV-2 S1/S2 IgG and Roche Elecsys Anti-SARS-CoV-2) versus results of molecular testing are shown in figure 1.

**Figure 1.**
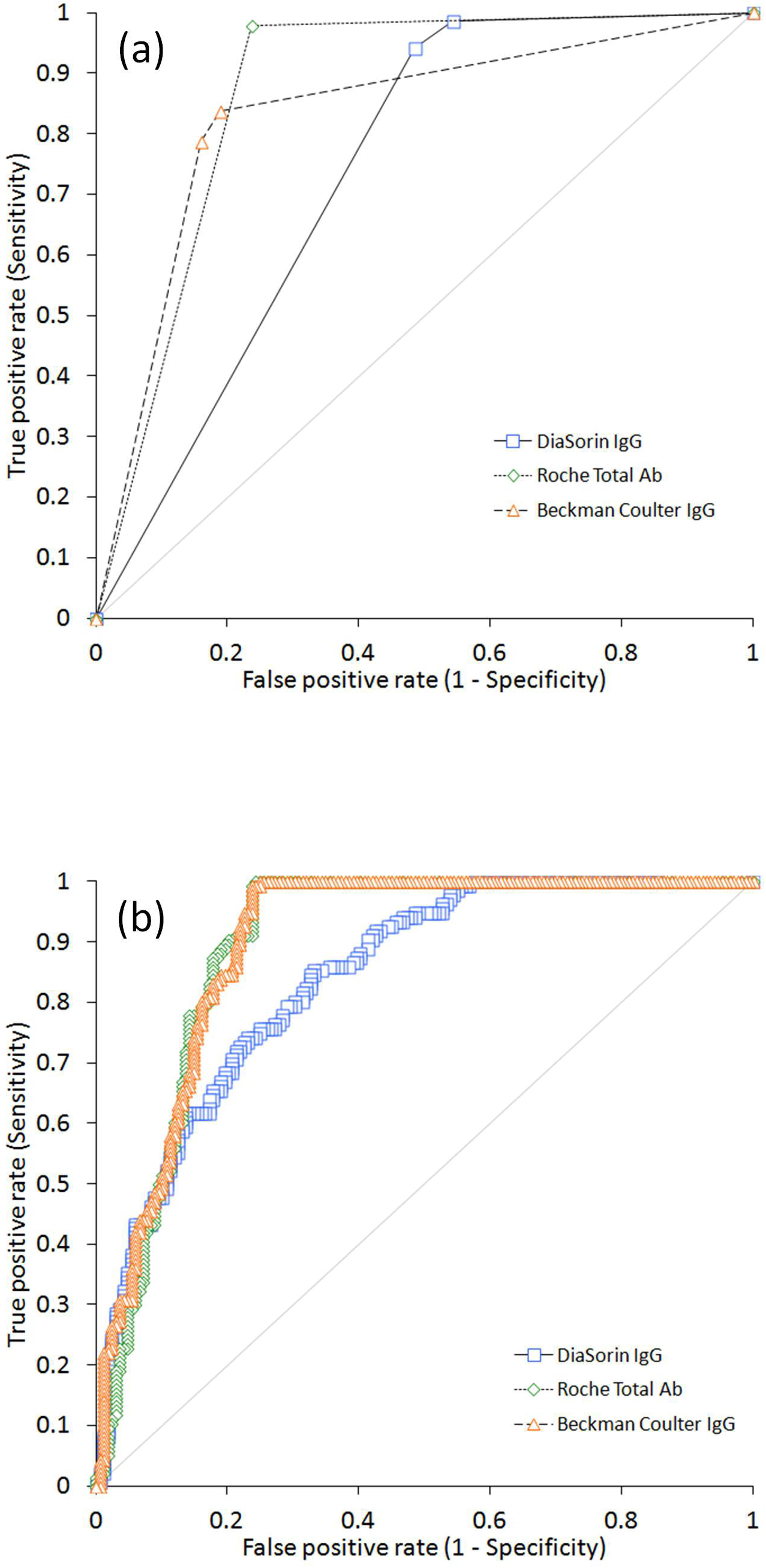
Diagnostic performance compared to molecular testing on nasopharyngeal samples of the three immunoassays used in this study. The area under the curve has been calculated using either (a) manufacturers’ cut-offs or (b) antibody titers.

As concerns the diagnostic accuracy using manufacturers’ cut-off (Figure 1a), Roche Elecsys Anti-SARS-CoV-2 displayed the highest AUC (0.87; 95% CI, 0.84-0.91; p<0.001), followed by Beckman Coulter Access SARS-CoV-2 IgG (0.83; 95% CI, 0.79-0.88; p<0.001) and DiaSorin Liaison SARS-CoV-2 S1/S2 IgG (0.74; 95% CI, 0.70-0.78). Notably, the AUCs of Beckman Coulter Access SARS-CoV-2 IgG and Roche Elecsys Anti-SARS-CoV-2 were not found to be statistically different (p=0.06), but were both significantly higher than that of DiaSorin Liaison SARS-CoV-2 S1/S2 IgG (both p<0.001). The diagnostic sensitivity at manufacturers’ cut-off (Table 1) was 0.98 for Roche Elecsys Anti-SARS-CoV-2, 0.94 for DiaSorin Liaison SARS-CoV-2 S1/S2 IgG and 0.79 for Beckman Coulter Access SARS-CoV-2 IgG, whilst the diagnostic specificity was 0.84 for Beckman Coulter Access SARS-CoV-2 IgG, 0.76 for Roche Elecsys Anti-SARS-CoV-2 and 0.52 for DiaSorin Liaison SARS-CoV-2 S1/S2 IgG, respectively.

The diagnostic accuracy using antibody titers is shown in figure 1b. The AUCs of all the three immunoassays appeared significantly improved, with Roche Elecsys Anti-SARS-CoV-2 (0.90; 95% CI, 0.86-0.93; p<0.001) and Beckman Coulter Access SARS-CoV-2 IgG (0.90; 95% CI, 0.86-0.94; p<0.001) displaying almost identical performance, and thus outstripping the diagnostic accuracy of DiaSorin Liaison SARS-CoV-2 S1/S2 IgG (0.84; 95% CI, 0.80-0.88). As for the diagnostic performance calculated using manufacturers’ cut-off, the AUCs of Beckman Coulter Access SARS-CoV-2 IgG (p<0.001) and Roche Elecsys Anti-SARS-CoV-2 (p=0.01) antibodies titers were found to be both significantly higher than that of DiaSorin Liaison SARS-CoV-2 S1/S2 IgG, whilst they were not significantly different between them (p=0.785). According to these results, the newly calculated diagnostic cut-offs, displaying the best performance for each assay, were as follows: 0.17 for Beckman Coulter Access SARS-CoV-2 IgG (1.00 sensitivity and 0.75 specificity); 0.36 for Roche Elecsys Anti-SARS-CoV-2 (1.00 sensitivity and 0.76 specificity); 22.9 for DiaSorin Liaison SARS-CoV-2 S1/S2 IgG (0.85 sensitivity and 0.67 specificity).

The Spearman’s correlation among the three different anti-SARS-CoV-2 CLIAs is shown in figure 2. The correlation of Beckman Coulter Access SARS-CoV-2 IgG was 0.80 (95% CI, 0.75-0.84; p<0.001) with Roche Elecsys Anti-SARS-CoV-2 and 0.72 (95% CI, 0.66-0.77; p<0.001) with DiaSorin Liaison SARS-CoV-2 S1/S2 IgG, respectively, whilst was 0.68 (95% CI, 0.61-0.73; p<0.001) between Roche Elecsys Anti-SARS-CoV-2 and DiaSorin Liaison SARS-CoV-2 S1/S2 IgG.

**Figure 2.**
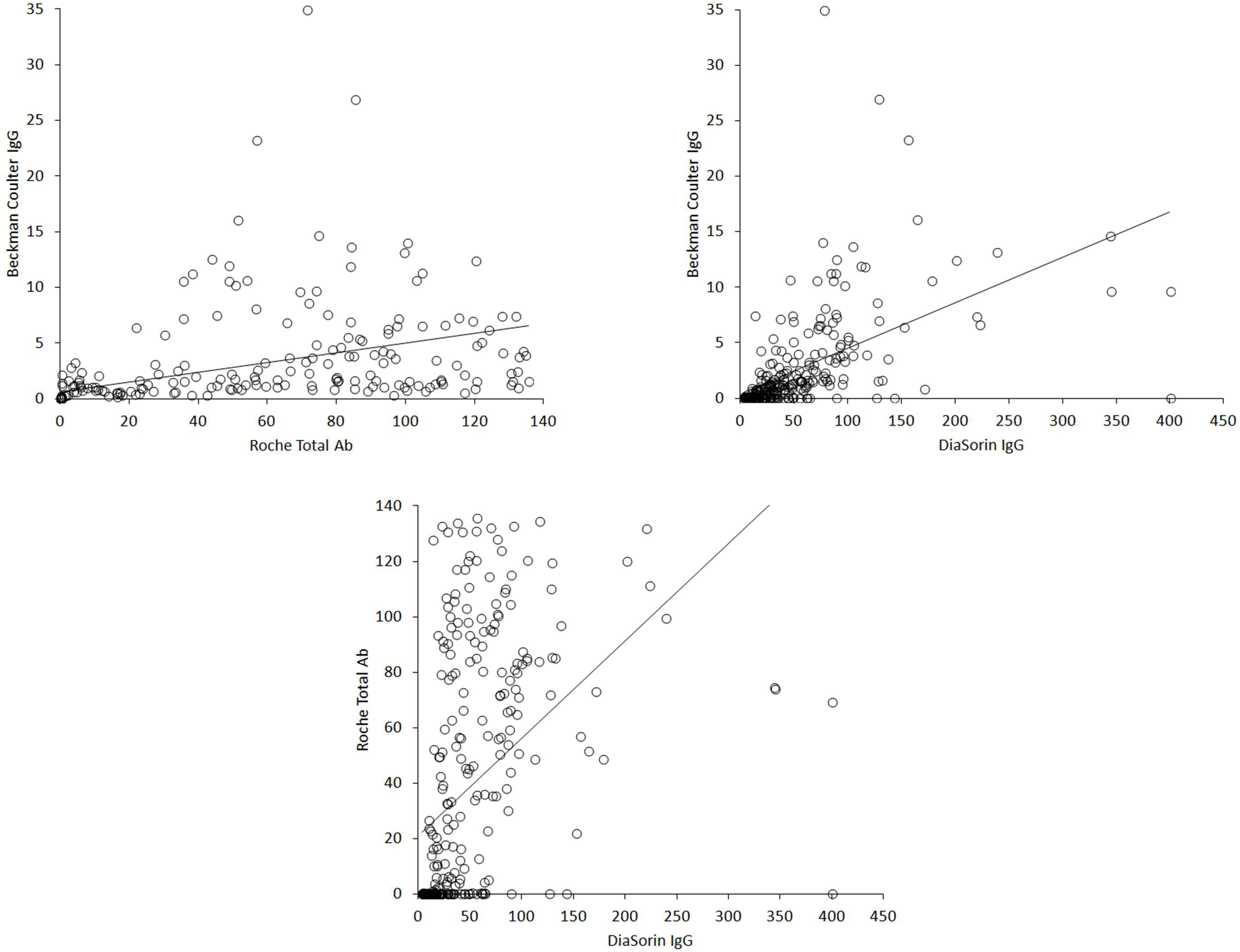
Spearman’s correlation among the three immunoassays used in this study.

## Discussion

Serological and seroprevalence surveys are considered an essential aspect for the clinical, economical and societal management of the ongoing COVID-19 pandemic outbreak (16). The identification of a humoral immune response developed against the target pathogen (i.e., SARS-CoV-2) is indeed the most important information that can be garnered from this type of testing, and that can then be straightforwardly used for establishing seroprevalence in specific geographies, environments or settings (i.e., communities, healthcare facilities, schools and so forth) (17), for studying nature, progression and duration of herd immunity (either natural or artificial) (18), for monitoring disease progression (19), as well as for complementing nucleic acid amplification testing (NAAT) under specific circumstances (20). Despite these important aspects, evidence remains that the analytical and diagnostic performance of many of marketed anti-SARS-CoV-2 tests is still limited, remains poorly validated, or even completely untested, thus contributing to raise serious doubts on their real clinical usefulness (21). Although high-quality serological assays are hence increasingly developed by many manufacturers all around the world, their implementation in laboratories of all size and nature needs to be anticipated by extensive validation of their analytical and clinical features, that would hence define their precise setting within a COVID-19 diagnostic pathway.

The novel Beckman Coulter Access SARS-CoV-2 IgG immunoassay has been developed for being used within routine workflow, and thus assisting clinical laboratories of virtually all dimensions and types for screening the presence of humoral (IgG) immune response against the virus in patients, healthcare professional and even in large community populations. Its suitability for full automation, characterized by random accessibility, contained turnaround time and high throughput, will enable rapid responses to massive testing programs, so enormously enhancing the efficiency of laboratory diagnostics within the context of the ongoing COVID-19 pandemics.

Our assessment of the analytical performance of this novel fully-automated CLIA has revealed a good repeatability profile, with total imprecision lower than ∼6%, a value that seems aligned with, or even better than, that reported in published evaluations of other commercially available anti-SARS-CoV-2 CLIAs (22-25). The linearity profile was also found to be optimal, in a range of IgG antibody titers between 0.11 and over 18, which is an upper limit of interval covering the vast majority of patient samples tested in the present study (302/305; >98%). The LoB, LoD and functional sensitivity were also found to be excellent, with a functional sensitivity that would enable to obtain a clinically usable antibody titers in most patients with ongoing or previous SARS-CoV-2 infection.

As concerns the diagnostic performance of Beckman Coulter Access anti-SARS-CoV-2 IgG immunoassay, the AUC of this method was found to be non-significantly different from that of Roche Elecsys Anti-SARS-CoV-2, an immunoassay specifically aimed at measuring total anti-SARS-CoV-2 antibodies, but definitely better than the AUC of DiaSorin Liaison SARS-CoV-2 S1/S2 IgG. It is hence conceivable that Beckman Coulter Access anti-SARS-CoV-2 IgG could be reliably used as surrogate of total antibodies within seroprevalence studies. This is not surprising since an abrupt and relatively rapid decline of anti-SARS-CoV-2 IgM and even IgA antibodies has been clearly described in patients with COVID-19 (26), thus making the assessment of antibodies classes other than IgG questionable (or even misleading) when performed weeks or months after symptoms relief. Nonetheless, unlike Roche Elecsys Anti-SARS-CoV-2 which targets the viral nucleocapsid protein, the Beckman Coulter Access anti-SARS-CoV-2 IgG CLIA has been developed against the spike protein and, more specifically, against the RBD. This would inherently mean that IgG quantification with this method would better mirror neutralizing activity compared to other immunoassays developed against different antigenic domains of SARS-CoV-2. Interestingly, our data are in keeping with those recently published by Tan et al., who also found an AUC of 0.947 in samples collected 21-64 days from symptom onset (27). Although we found that Beckman Coulter Access anti-SARS-CoV-2 IgG immunoassay exhibited a slightly lower diagnostic sensitivity compared to that reported by Chua et al using the manufacturer’s cut-off (i.e., 0.79 vs. 0.85) (28), local recalculation of the diagnostic threshold was effective to enhance the diagnostic sensitivity to 1.00 (vs. 0.79), while only slightly affecting the diagnostic specificity (i.e., 0.84 vs. 0.75), thus confirming previous evidence published by on this matter (19).

The considerably lower value of the diagnostic cut-offs of both Roche Elecsys Anti-SARS-CoV-2 total antibodies and Beckman Coulter Access anti-SARS-CoV-2 IgG, as recalculated from the locally generated ROC curves, deserves a distinct scrutiny. The manufacturer-suggested diagnostic thresholds are typically derived for optimizing the diagnostic performance of the assay during acute SARS-CoV-2 infections, when antibody titers are the highest (29). However, since the circulating levels of all anti-SARS-CoV-2 Ig classes tend to gradually decline over time, this arbitrary cut-offs would become predictably too high to retain the same diagnostic performance over a long period of time after symproms relief. It is hence conceivable that multiple diagnostic thresholds would need to be identified for all anti-SARS-CoV-2 immunoassays, according to the time passed from symptoms onset. This conclusion is reinforced by the evidence garnered in our study, since the diagnostic sensitivity of Roche Elecsys Anti-SARS-CoV-2 and Beckman Coulter Access anti-SARS-CoV-2 IgG in samples collected up to 4 months after achieving a molecular diagnosis of SASR-CoV-2 infection improved from 0.98 and 0.79 up to 1.00 for both immunoassays when the cut-offs values were lowered by 83% and 64%, respectively. Failing to do so would be associated with a progressive decrease of the diagnostic sensitivity (i.e., higher rate of false negative values), which may then lead to the underdiagnosis of a substantial number of patients who were instead really infected by the virus.

## Conclusions

Although several progresses have been made in our current understanding of COVID-19, there is still more to learn on biochemistry, biology and clinics of SARS-CoV-2 infection (30). Laboratory diagnostics is not an exception to this rule, whereby the ideal usage of SARS-CoV-2 serology remains a challenging enterprise. The results of our analytical evaluation of Beckman Coulter Access anti-SARS-CoV-2 IgG immunoassay supports the conclusion that this fully-automated CLIA represents a valuable resource for large and accurate seroprevalence surveys. Further studies would then be needed to clarify many undefined issues in anti-SARS-CoV-2 antibody testing.

## Data Availability

Data are availabile from the authors upon request.

## Footnote

## Ethical Statement

The authors are accountable for all aspects of the work in ensuring that questions related to the accuracy or integrity of any part of the work are appropriately investigated and resolved. This is a retrospective investigation, based on pre-existing patient samples, collected for routine SARS-CoV-2 testing, and thereby no patient’s informed consent or Ethical Committee approval were necessary.

## Open Access Statement

This is an Open Access article distributed in accordance with the Creative Commons Attribution-NonCommercial-NoDerivs 4.0 International License (CC BY-NC-ND 4.0), which permits the noncommercial replication and distribution of the article with the strict proviso that no changes or edits are made and the original work is properly cited (including links to both the formal publication through the relevant DOI and the license). See: https://creativecommons.org/licenses/by-nc-nd/4.0/.

